# A method for estimating causal effects from heterogeneous clinical trials without a common control group using sequential regression and simulation: an individual participant data meta-analysis and validation study

**DOI:** 10.1101/2022.10.07.22280801

**Authors:** Vivek A. Rudrapatna, Vignesh G. Ravindranath, Douglas V. Arneson, Arman Mosenia, Atul J. Butte, Shan Wang

## Abstract

**BACKGROUND:** The advent of clinical trial data sharing platforms has created opportunities for making new discoveries and answering important questions using already collected data. However, existing methods for meta-analyzing these data require the presence of shared control groups across studies, significantly limiting the number of questions that can be confidently addressed. We sought to develop a method for meta-analyzing potentially heterogeneous clinical trials even in the absence of a common control group.

**METHODS:** This work was conducted within the context of a broader effort to study comparative efficacy in Crohn’s disease. Following a search of clnicaltrials.gov we obtained access to the individual participant data from nine trials of FDA-approved treatments in Crohn’s Disease (N=3392). We developed a method involving sequences of regression and simulation to separately model the placebo- and drug-attributable effects, and to simulate head-to-head trials against an appropriately normalized background. We validated this method by comparing the outcome of a simulated trial comparing the efficacies of adalimumab and ustekinumab against the recently published results of SEAVUE, an actual head-to-head trial of these drugs. This study was pre-registered on PROSPERO (#157827) prior to the completion of SEAVUE.

**FINDINGS:** Using our method of sequential regression and simulation, we compared the week eight outcomes of two virtual cohorts subject to the same patient selection criteria as SEAVUE and treated with adalimumab or ustekinumab. Our primary analysis replicated the corresponding published results from SEAVUE (p=0·9). This finding proved stable under multiple sensitivity analyses.

**INTERPRETATION:** This new method may help reduce the bias of individual participant data meta-analyses, expand the scope of what can be learned from these already-collected data, and reduce the costs of obtaining high-quality evidence to guide patient care.

**FUNDING:** NIH; UCSF Division of Gastroenterology and Bakar Computational Health Sciences Institute; University of San Francisco

## INTRODUCTION

The individual participant data (IPD) meta-analysis is the gold-standard for clinical research^1,2^. Access to the raw data from trials affords investigators the opportunity to verify published results, ask new questions of these data, and uncover findings with the potential to impact patient care.

Performing an IPD meta-analyses usually requires multiple trials with negligible heterogeneity across many dimensions: cohort definition, randomization, blinding, parallel study arms, interventions, and outcomes^1,2^. Although this requirement ensures unbiased estimation, it substantially limits the number of meta-analyses that can be performed due to the rarity of replicate trials.

The method of mixed-effects regression is commonly used to address study heterogeneity when meta-analyzed trials include a shared control group (i.e. placebo). However, there is a paucity of methods for common situations where there is no shared control group across potential studies. The few methods that have been developed include naïve pooling^3^, as well as the Bayesian method of power priors^4–7^. However, these methods fail to address the problem of cohort heterogeneity^8,9^. Another major limitation is the lack of external validation against prospective studies. The result of this methodological gap is the common practice of excluding uncontrolled studies from potential meta-analyses, and ultimately fewer research questions that we are statistically powered to answer using already-collected data.

Here we report a new method for meta-analyzing clinical trials data in the absence of a common control group. We illustrate our method of sequential regression and simulation in the context of a comparative efficacy analysis in Crohn’s disease, an immune disorder of the gastrointestinal tract. We use the data from six placebo-controlled trials (N=3153) to develop a model of the placebo effect, then apply this to three placebo-less trials (N=239) to normalize and separately model the drug-attributable response. Finally, we validate the method by predicting the results of SEAVUE (NCT03464136), a recent head-to-head trial of ustekinumab versus adalimumab^10^.

## METHODS

This study was approved by the UCSF IRB (#18-24588). It was pre-registered on PROSPERO^11^ (#157827), YODA^12^, and Vivli^13^ prior to the initiation of this work and the completion of SEAVUE.

### DATA ACCESS

In June 2019 we queried clinicaltrials.gov to identify studies for meta-analysis (Supplemental Data: Figure 1, Table 1). We found 90 trials listed as completed, phase 2-4, randomized, double-blinded, interventional trials of FDA-approved treatments for Crohn’s disease. We manually confirmed 16 trials as meeting these criteria. To ensure comparability of these studies, we reviewed their major inclusion and exclusion criteria and confirmed that the Crohn’s Disease Activity Index (CDAI) had been captured at week six or eight relative to treatment initiation. We confirm that these studies were at low risk of bias using the Cochrane ROB2 tool. We obtained access to the IPD for 15 studies (N=5703). They were conducted between 1999 and 2015 and corresponded to all six FDA-approved biologics as of 2019.

**Table 1:**
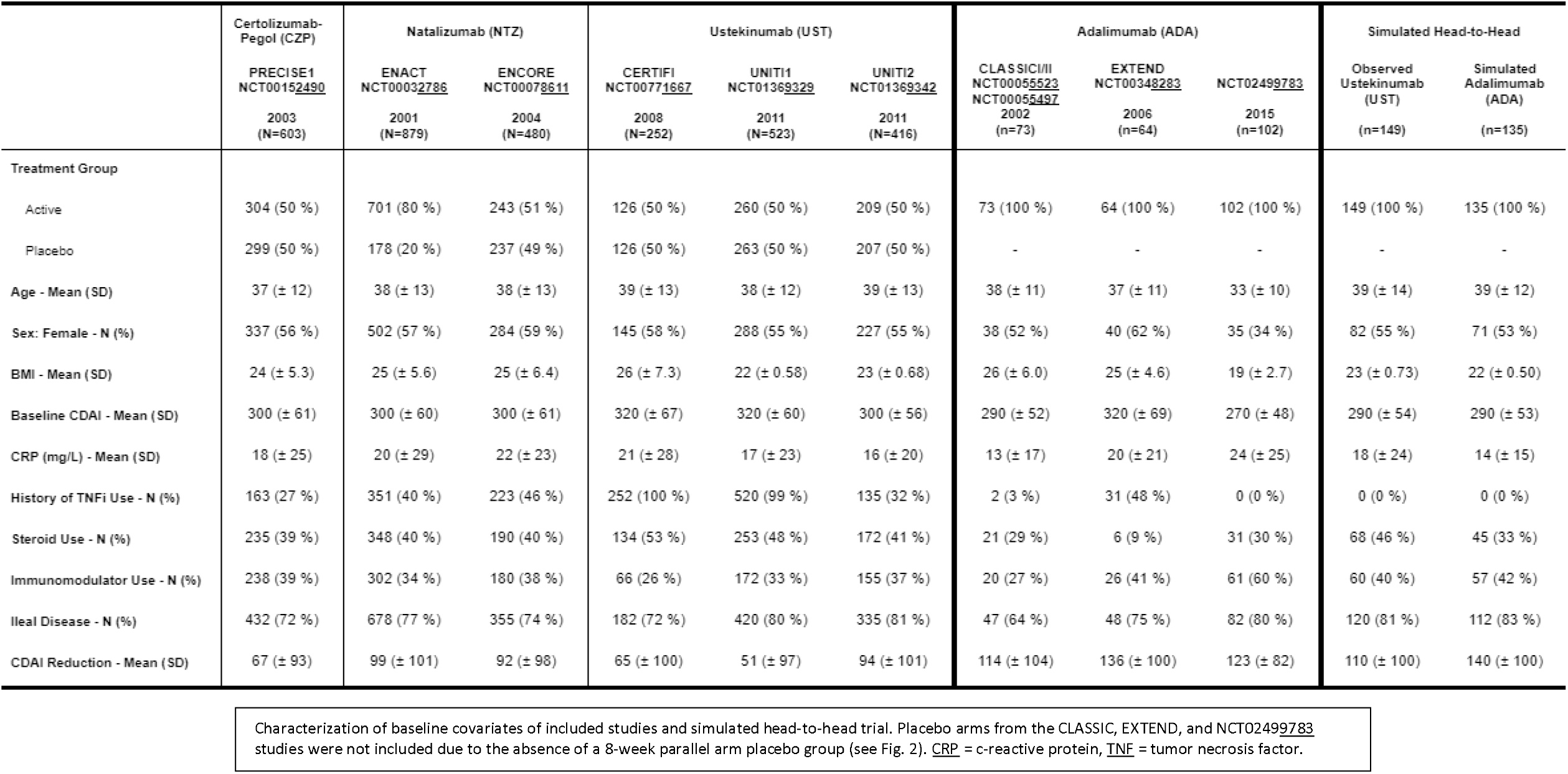
Characterization of included studies.

**Figure 1:**
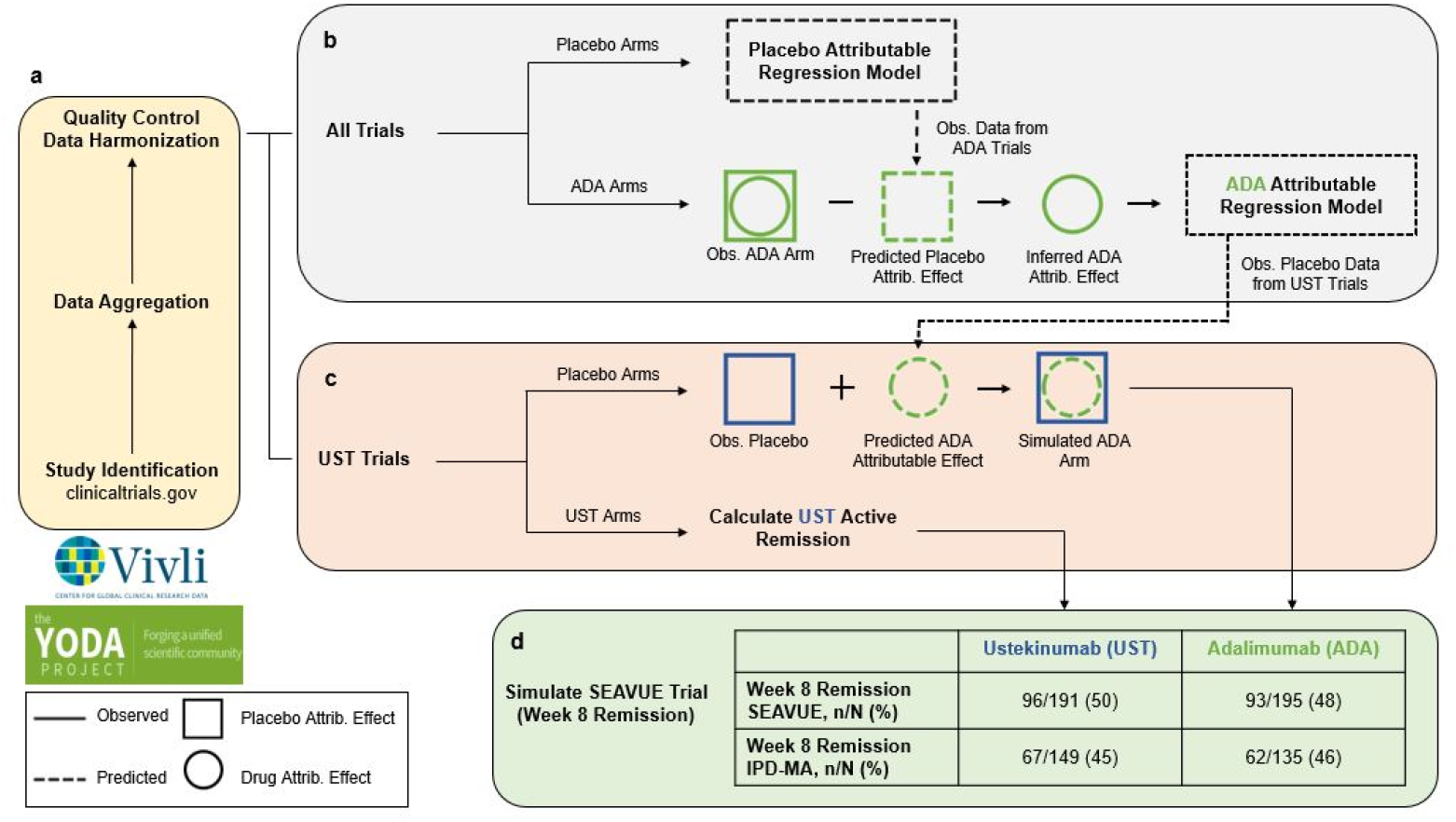
Overview of modeling process. Overview of creating a simulated head-to-head trial of ustekinumab (UST; blue) and adalimumab (ADA; green) in the absence of a common control group. **a**, Clinical trials were found using clinicaltrials.gov and sought for retrieval on the YODA and Vivli platforms. Individual participant data (IPD) from trials that collected CDAI scores at week 8 visits were then aggregated and harmonized. **b**, Two linear mixed effect models - placebo-attributable and ADA-attributable - were developed from the harmonized data to partition the CDAI reduction a participant experience based on baseline covariates (age, sex, BMI, etc.). Disease reduction was partitioned into placebo attributable (square) and drug attributable (circle) effects; IPD (solid lines) were used to predict or simulate data (dashed lines). **c**, Using the ADA-attributable model, a simulated ADA arm was developed. **d**, Results from a simulated head-to-head trial were compared against a recently completed head-to-head trial, SEAVUE, to externally validate the proposed method.

### STUDY DESIGN

We designed this study to emulate a hypothetical head-to-head, parallel-design, efficacy trial randomizing participants to two treatment arms (Figure 1). Although a typical meta-analytic study design would have involved pooling cohorts from several internally controlled, parallel-arm trials, this was not possible in our case for several reasons. For example, many studies involved open-label induction followed by a randomization event to continue or discontinue the treatment (Figure 2).

**Figure 2:**
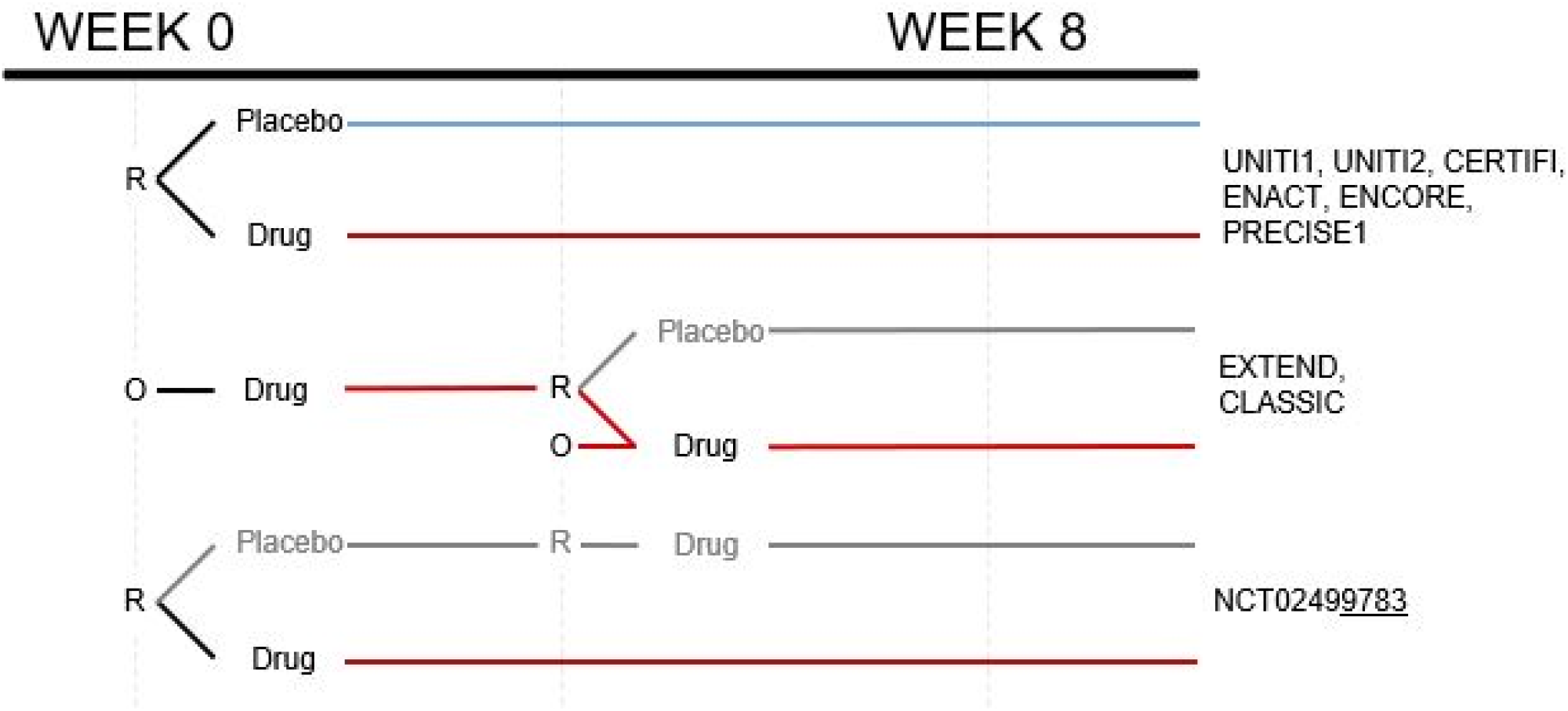
Summary of randomized controlled trial study designs. Data harmonization required careful understanding of the study designs. All treatment arms that involved 8 weeks of consistent exposure to either placebo or (blue) or active treatment at the FDA-approved doses (red) were included. R = randomized and blinded; O = open label.

To overcome this heterogeneity, we defined our primary outcome as the absolute reduction in CDAI at week eight, and we filtered the provided data to include only those cohorts that had at least eight weeks of uninterrupted observation time on either placebo or a drug relative to baseline. We noted that nine of the 15 trials did not include a parallel arm placebo cohort randomized at week 0 and followed for eight weeks. Thus, for our study, they were considered uncontrolled. As a first step towards developing a method for handling this heterogeneity, we restricted our initial analyses to just the placebo-controlled trials (six trials; N=3153; Table 1). Subsequent analyses used a second set of placebo-less trials of adalimumab, one of the drugs compared in SEAVUE.

**Supplemental Figure 1:**
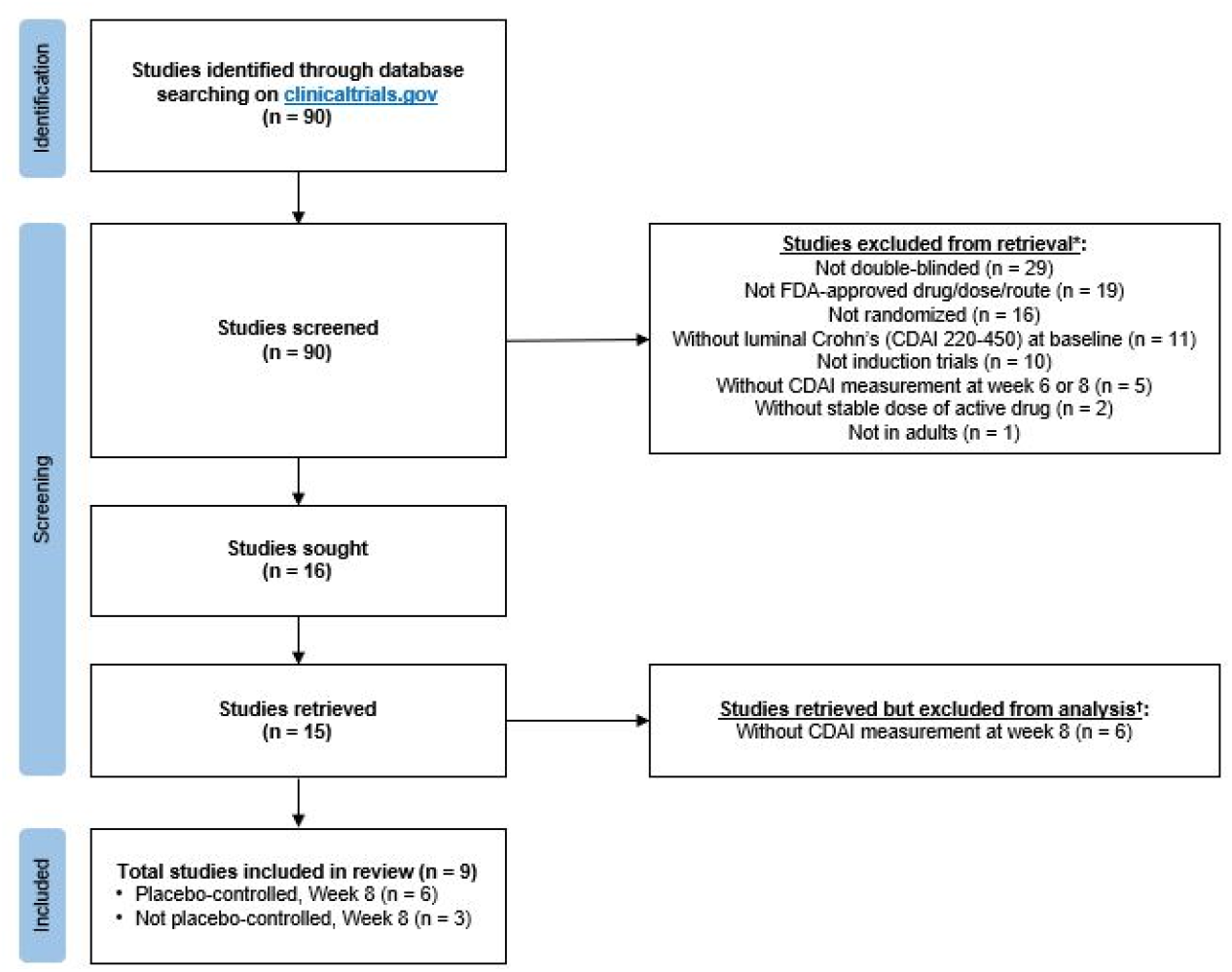
PRISMA-IPD flow diagram. Flow diagram illustrating selection of studies. *Some studies met more than one criterion. †All 15 studies were retrieved and consolidated on the Vivli platform; however, only 9 studies were used for analysis as these studies captured CDAI measurement at week 8 and could be compared with the SEAVUE study.

**Supplemental Table 1:** Major inclusion/exclusion criteria of included studies.

| Trial | Age ≥ 18 | Baseline CDAI 220-450 | Ileal and/or colonic involvement | Disease activity by biochemical/imaging/endoscopy | TNFi intolerance/failure | TNFi naive* | Stable concomitant medications | No symptomatic stricture | No abscess | No recent surgery | No stoma or ostomy |
| --- | --- | --- | --- | --- | --- | --- | --- | --- | --- | --- | --- |
| PRECISE1 | ✓ | ✓ | NA | NA | ✗ | ✓ | ✓ | ✓ | ✓ | NA | ✓ |
| ENACT | ✓ | ✓ | ✓ | ✓ | ✗ | ✗ | ✓ | ✓ | ✓ | NA | ✓ |
| ENCORE | ✓ | ✓ | NA | ✓ | ✗ | ✗ | ✓ | ✓ | ✓ | ✗ | ✓ |
| CERTIFI | ✓ | ✓ | ✓ | ✗ | ✓ | ✗ | ✓ | ✓ | ✓ | ✓ | ✓ |
| UNITI1 | ✓ | ✓ | ✓ | ✗ | ✓ | ✗ | ✓ | ✓ | ✓ | ✓ | ✓ |
| UNITI2 | ✓ | ✓ | ✓ | ✓ | ✗ | ✓ | ✓ | ✓ | ✓ | ✓ | ✓ |
| CLASSIC | ✓ | ✓ | NA | NA | ✗ | ✓ | ✓ | ✓ | NA | ✓ | ✓ |
| EXTEND | ✓ | ✓ | ✓ | ✓ | ✗ | ✗ | ✓ | NA | NA | NA | NA |
| NCT02499783 | ✓ | ✓ | ✗ | ✓ | ✗ | ✓ | ✓ | ✓ | ✗ | ✓ | ✓ |

If the trial protocol was not publicly available, and if the corresponding manuscript or clinical study report was silent on a given criteria, the field was annotated as NA. If the trial protocol was available and if it was clear that a given criterion was not applied for cohort selection, the field was annotated with an X. In many trials (e.g., ENACT, ENCORE), a history of TNF-naïve or intolerance/failure was not a requirement and was captured as a participant-specific covariate for regression-based control. In other scenarios, the trial-specific covariate implicitly applied to all trial participants (e.g., TNF-naïve status in PRECISE1). Trials were generally consistent on the target patients of study in terms of inclusion and exclusion criteria. To address the possibility of residual heterogeneity due to the lack of perfectly consistent eligibility criteria or unmeasured covariates, a trial-specific random effect was included in the final regression models.

**Supplemental Figure 2:**
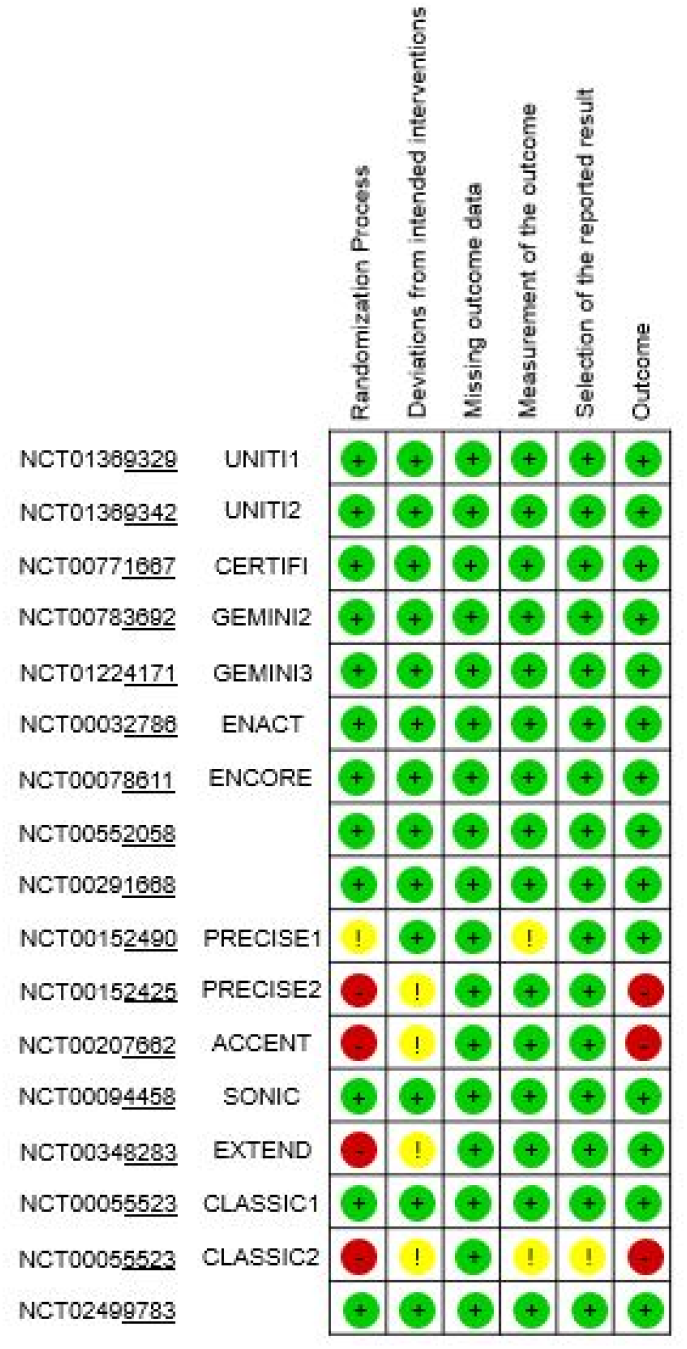
Risk of Bias. Cochrane’s risk-of-bias tool for randomized trials version 2 (ROB2). Green, yellow, and red indicate low, moderate, and high risk of bias respectively.

### QUALITY CONTROL, HARMONIZATION

We performed extensive tests of data quality (Supplemental Methods). These included reproducing published results from each trial cohort (Supplemental Figures 3, 4). We used domain knowledge to select nine variables that were universally available across trials for subsequent modeling: Age, Sex, body mass index (BMI), baseline CDAI, c-reactive protein (CRP), history of tumor necrosis factor-alpha inhibitor (TNFi) use, steroid use, immunomodulator use, and ileal involvement.

3% of the participants had at least one missing covariate at baseline. Continuous variables were addressed by median imputation, and participants with missing categorical variables were dropped (N=86). 11% of participants had a missing outcome at week eight. We used last-observation-carried-forward to impute these. This is the typical practice for the analysis of these trials in regulatory submissions and was the prespecified approach in the protocols of the included trials.

Some important variables could not be included in this study. Ethnicity was not collected in most trials. Race was missing in some trials, but when it was captured, it reflected significant imbalance (88% of participants were white). Other variables like disease behavior and duration were not uniformly captured across studies.

### MODELING ASSUMPTIONS

We incorporated several assumptions when developing and interpreting candidate models. We assumed that the observed week eight reduction in CDAI reflected a combination of two distinct effects: a drug-independent (i.e., placebo) effect and drug-attributable effect. These effects were separately modeled as a function of the above predictors and study year. The justification for this is briefly summarized below and additionally presented graphically (Figure 3).

**Figure 3:**
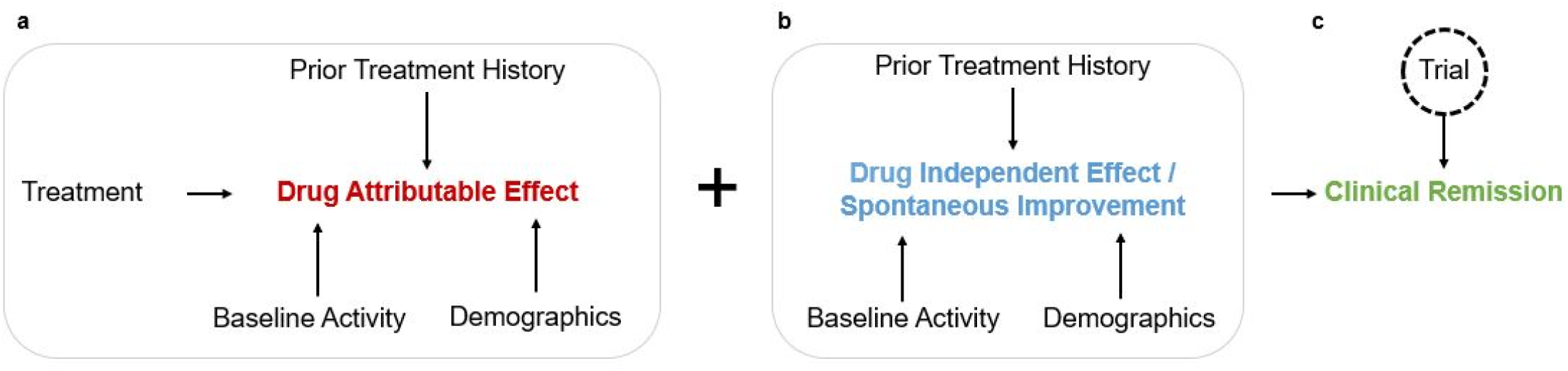
Directed acyclic graph. **a**, A directed acyclic graph (DAG) of the drug attributable effect. In addition to the active treatment itself, patient demographics (e.g. age, sex, BMI), baseline Crohn’s disease activity (e.g. baseline CDAI, CRP, location), and treatment history (e.g. prior use of TNFis, current use of oral corticosteroids and immunomodulators) are all modelled as contributing to the drug attributable effect. The non-drug covariates are effect modifiers and are implicitly modeled as two-way interaction terms with the active drug. **b**, A DAG of the drug independent effect (i.e. placebo effect). The same covariates except for the treatment term are modeled as effect modifiers and are implicitly represented as two-way interactions with the placebo effect. **c**, Drug attributable and drug independent effects have additive effects on the overall clinical remission at week 8 (CDAI < 150), with any individual trial reflecting a noisy measurement of the true effect due to unmodeled heterogeneity in study design and execution (random effect).

The placebo effect was modeled as a function of the nine covariates as well as predictors of trial-specific heterogeneity. We assumed that much of the spontaneous improvement seen in placebo-assigned participants was related to regression to the mean, as study participation was limited to patients with currently active Crohn’s disease. Conversely, we assumed that failure to spontaneously improve was likely to reflect chronic and cumulative disease burden with relative stability in symptoms. Thus, variables corresponding to concomitant and prior treatments were treated as proxies of chronic disease burden and included as predictors. Lastly, we considered other influences on overall heterogeneity, including differences in cohorts, data capture, outcome ascertainment, and study personnel. To account for these sources of variation, we included study year as well as trial identifier as additional covariates. In mixed-effect models, trial was included as a random effect. Other covariates were fixed effects.

The drug-attributable effect was separately modeled as a function of these same covariates, reflecting drug-specific (interaction) effects on the outcome. Many of these covariates are well-established as modifiers of treatment response, such as a history of TNFi and immunomodulators use^4^. Others (CRP, baseline CDAI) are proxies of bowel inflammation, the target of these medications. These variables were included to maximize the explained variation in the outcome.

### DEVELOPMENT AND ASSESSMENT OF A PLACEBO MODEL

We fit a linear mixed effects model utilizing all nine predictors as well as study year as predictors of the placebo effect. To minimize the risks of residual bias due to model misspecification (e.g., non-linearities, unmodeled interactions), we compared the predictive performance of this model against other statistical and machine learning models. We further evaluated this model from the perspective of being used to impute unmeasured placebo effects, and thus normalize different trials to the same background. We performed a leave-one-trial-out analysis and inspected the trial-averaged residuals.

**Supplemental Figure 3:**
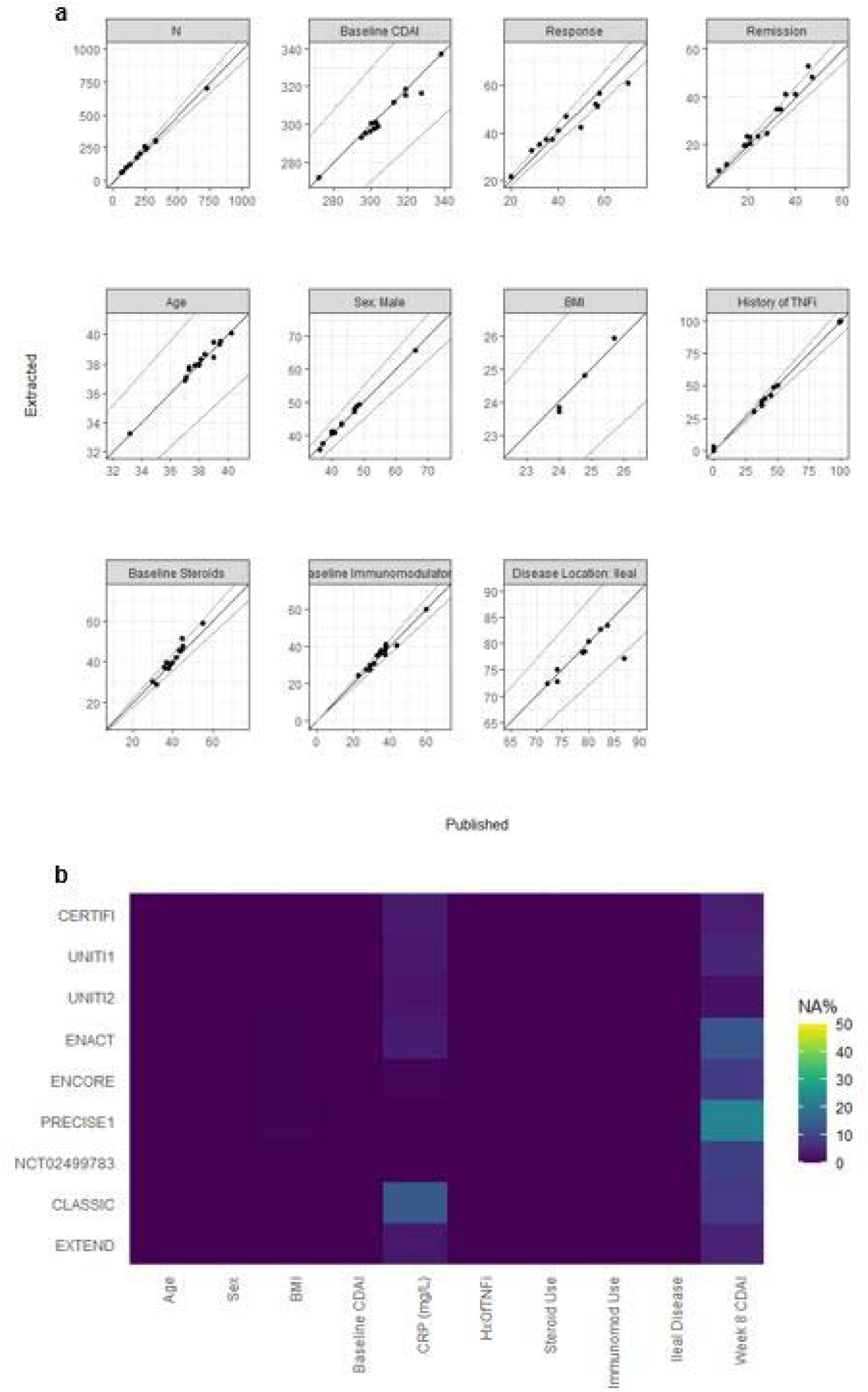
Reproducibility of published data. **a**, Plots of aggregated data versus published data for baseline covariates and outcome variables as a measure of quality control. Each dot represents the mean variable estimate for a given study treatment group (placebo, active). Data were not displayed if the study did not report the variable mean in its original article. Upper and lower lines in plots correspond to ±10% error bounds. **b**, Percentage of missing covariates by study. Approximately 0.2% of BMI values, 2% of CRP values, and 11% of week 8 CDAI values were missing after data harmonization and required imputation. Median imputation by study was used to impute missing BMI and CRP values. Last observation carried forward (LOCF) was used to impute missing week 8 CDAI values; CDAI observations from week 6, week 4, week 3, or week 2 were candidates for LOCF.

**Supplemental Figure 4:**
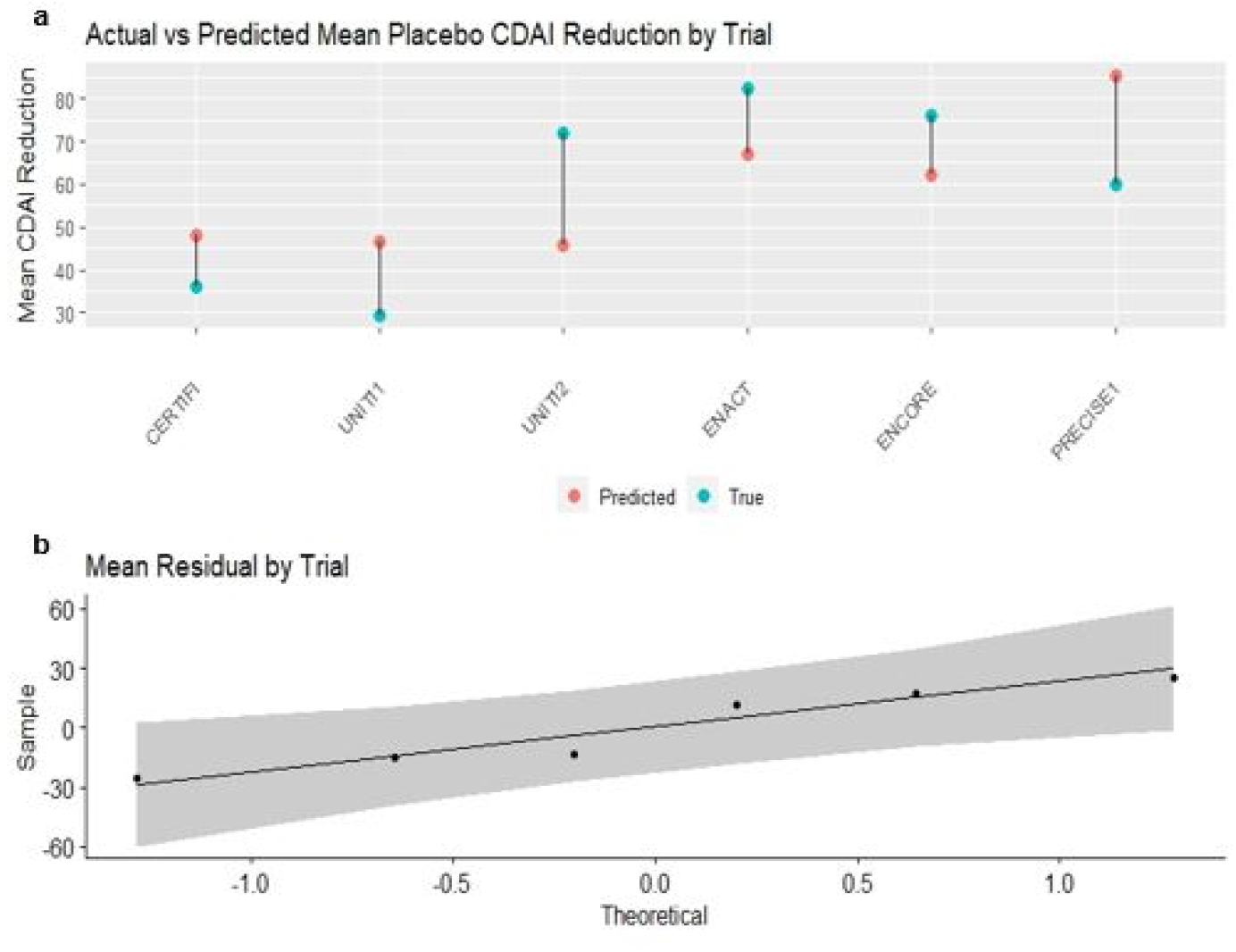
Leave-one-trial-out analysis. **a**, A leave-one-trial-out analysis, where blue and red dots represent the true and predicted mean CDAI reduction respectively. **b**, Q-Q plot of the model residuals (difference in true and predicted mean CDAI reduction values per study) to assess residual normality (p=0.4 by the Shapiro-Wilk test).

**Supplemental Figure 5:**
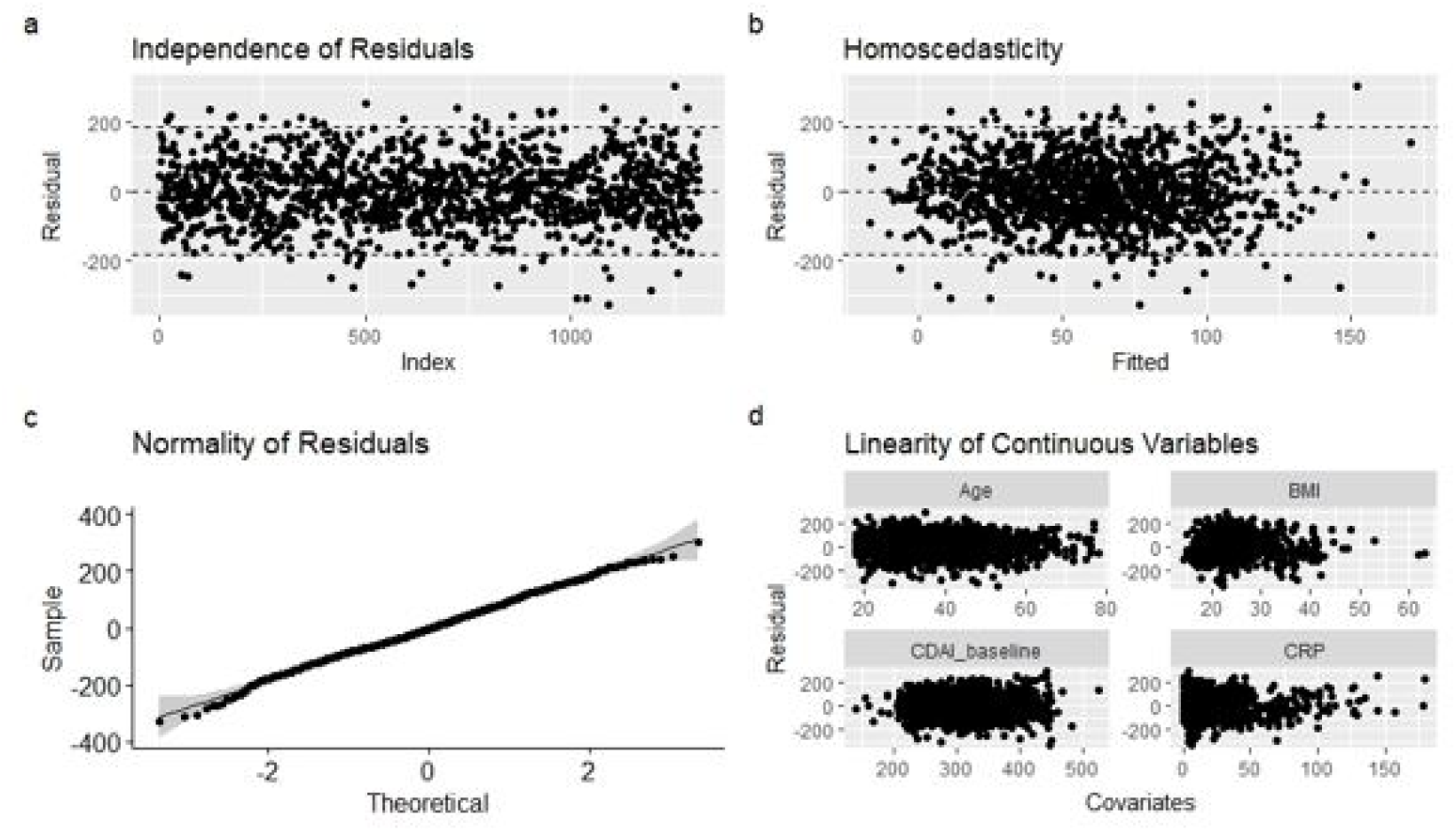
Checking model assumptions. Plotting the placebo-attributable model residuals to visually check linear regression model assumptions. **a**, Plot of the model residuals versus index to assess residual independence. Dotted lines represent ±2**σ. b**, Plot of the model residuals versus the fitted values to assess residual homoscedasticity. **c**, Q-Q plot of the model residuals to assess residual normality. **d**, Plot of the model residuals versus each continuous covariate to assess linearity between covariates and the outcome variable.

### ESTIMATION OF THE DRUG-ATTRIBUTABLE EFFECT

To normalize the responses of drug-assigned cohorts that lacked a within-study, parallel-arm control group, we used the finalized placebo model to simulate and subsequently partition their overall response into drug-independent and drug-dependent (i.e., drug attributable) components (Figure 1b). We performed this using the data from the adalimumab trials (N=239) because they were all lacking an 8-week continuous placebo group and thus required normalization. From the outcomes of these patients, we subtracted the conditional mean outcomes associated with the placebo effect and used the residuals as the new outcome variable of a second mixed-effects model to estimate the adalimumab-attributable effect.

### VALIDATION, SENSITIVITY ANALYSES

Using the covariates associated with the placebo recipients of the ustekinumab trials, we used the adalimumab-attributable regression model to simulate their counterfactual week 8 outcomes had they received adalimumab instead (Figure 1c). We identified the subset of these virtual patients who were naïve to TNFi (an additional inclusion criterion from SEAVUE) and did the same with the ustekinumab recipients. We compared their week 8 outcomes using the same definition of clinical remission as used in SEAVUE (CDAI < 150), and performed a Fisher’s exact test to compare our results with SEAVUE’s. We tested the robustness of our result using three sensitivity analyses: 1) removing ENACT and ENCORE from the dataset due to >10% missingness of outcome data, 2) removing participants with missing outcomes, and 3) removing the ustekinumab trials from the placebo model training data, to address potential information leakage. Lastly, we compared our results with what we might have found had we not used our method to normalize cohorts.

## RESULTS

See Figure 1 for an overview. This method was originally developed in the context of an existing effort to study comparative efficacy in Crohn’s disease by reanalyzing the IPD of corresponding clinical trials. As the first step towards this goal, we sought to address the problem of meta-analyzing data from several potentially heterogeneous trials lacking a common control group.

### DATA ACCESS

We queried clinicaltrials.gov and performed manual review to confirm 16 trials as meeting these criteria: completed, phase 2-4, randomized, double-blinded, interventional trials of FDA-approved treatments for Crohn’s disease as of June 2019 (Figure 1a, Supplemental Figure 1). Included trials had common inclusion/exclusion criteria, or had participant-level data available to control for this heterogeneity (Supplemental Table 1). They all measured the same endpoint (CDAI) at week eight and were at low risk of bias (Supplemental Figure 2). We obtained access to the IPD for 15 studies (N=5703), corresponding to trials of all six FDA-approved biologics as of 2019.

### DEVELOPMENT AND ASSESSMENT OF A PLACEBO MODEL

We fit a linear mixed effects model utilizing nine clinical features and study year as predictors of the placebo effect (Figure 1b, Table 2). To minimize the risk of residual bias due to model misspecification, we compared the predictive performance of this model against other machine learning models (Supplemental Table 2). We found no significant differences in the root-mean-squared-error. Thus, we selected the mixed-effects model for downstream analyses.

**Table 2:** Mixed effect models.

| Predictors | Placebo Attributable Model<sup>a</sup> |  |  | ADA Attributable Model<sup>b</sup> |  |  |
| --- | --- | --- | --- | --- | --- | --- |
|  | Estimates | SE | p | Estimates | SE | p |
| Intercept | 92.18 | 13.12 | <0.001 | 67.57 | 20.41 | 0.001 |
| Year (Centered) | -1.89 | 1.55 | 0.223 | - | - | - |
| Baseline CDAI (Centered) | 0.37 | 0.04 | <0.001 | 0.12 | 0.11 | 0.276 |
| Age (Centered) | 0.13 | 0.21 | 0.549 | -1.77 | 0.57 | 0.002 |
| BMI (Centered) | 0.52 | 0.54 | 0.341 | -1.45 | 1.34 | 0.280 |
| CRP (mg/L) (Centered) | -0.28 | 0.11 | 0.012 | -0.35 | 0.28 | 0.213 |
| Sex: Male | -0.72 | 5.17 | 0.889 | 7.14 | 12.13 | 0.556 |
| History of TNFi Use | -37.56 | 6.32 | <0.001 | 7.51 | 20.38 | 0.712 |
| Steroid Use | 7.15 | 5.24 | 0.172 | 5.36 | 14.35 | 0.709 |
| Immunomodulator Use | 5.42 | 5.45 | 0.320 | -17.43 | 12.54 | 0.164 |
| Ileal Disease | -7.46 | 5.99 | 0.213 | 2.84 | 13.74 | 0.836 |

Mixed effect linear regression outputs for the placebo attributable (n=1310) and ADA attributable (n=239) models. For training, Year was centered by subtracting 2000, Baseline CDAI was centered by subtracting 300, Age was centered by subtracting 35, BMI was centered by subtracting 20, and CRP (mg/L) was centered by subtracting 10. ICC = intraclass correlation coefficient. **a.** The placebo attributable model (ICC 0.02) trial random intercepts were found to be PRECISE1: -12.808, UNIT1: -7.975, CERTIFI: -6.328, ENACT: 6.077, ENCORE: 8.669, and UNITI2: 12.366. **b.** ADA attributable model (ICC 0.05) trial random intercepts were found to be CLASSIC: -20.215, EXTEND: 9.439, and NCT02499783: 10.775.

We evaluated this model from the perspective of being used to impute unmeasured placebo effects, and thus normalize different trials to the same background placebo response. A leave-one-trial-out analysis suggested that the model predictions were robust and unbiased (Supplemental Figures 4, 5). The trial-averaged residuals were consistent with normality (p=0·4; Shapiro-Wilk test).

We noted that the unmodeled variation in the placebo effect was relatively large and was independent of the choice of model (Supplemental Table 2). These results explain the large placebo effects that have been seen in Crohn’s disease randomized trials (regression to the mean), and suggest that more work will be needed to improve the measurement of Crohn’s disease activity.

**Supplemental Table 2:** Model selection.

| Prediction Model | Package | Mean 5-Fold CV RMSE |
| --- | --- | --- |
| Linear Regression | lm | 93.19 |
| Linear Mixed Effect Model | lme4 | 93.19 |
| Linear LASSO Regression | glmnet | 93.14 |
| Random Forest | randomForest | 92.85 |
| XGBoost (DART) | xgboost | 96.20 |
| Stacked Ensemble | MLJAR | 90.81 |

Mean 5-fold cross-validation (CV) root mean squared error (RMSE) scores of various predictive models. All models were tested against the same stratified 5-fold datasets to ensure results were comparable. RMSE scores reflect the model's ability to predict CDAI reduction at week 8 due to a placebo treatment given baseline covariates (age, sex, BMI, etc.). The stacked ensemble model was built from 9 default base models using the mljar AutoML package.

To study the placebo effect and identify potential opportunities to improve trial efficiency, we reviewed all significant predictors. A history of TNFi was associated with a 38-point reduction in the placebo effect. We interpreted this as reflecting a greater cumulative disease burden in patients who failed to improve with TNFis, with disease complications (e.g., minor intestinal strictures) that are unlikely to spontaneously regress over 8 weeks. Similarly, CRP was a negative predictor, suggesting that untreated acute inflammation is unlikely to improve over short time periods. The baseline CDAI was a positive predictor, likely reflecting regression to the mean effects. Age, sex, BMI, concomitant medications, and ileal involvement were not found significant, potentially due to multicollinearity.

### ESTIMATION OF THE DRUG-ATTRIBUTABLE EFFECT

We sought to normalize the responses of drug-assigned cohorts that lacked a within-study, parallel-arm control group. Our strategy was to use the finalized placebo model to partition the overall response into drug-independent and drug-attributable components (Figure 1b). We applied this approach to the data from three study cohorts assigned to receive adalimumab at the FDA-approved dose for treatment induction (N=239; Table 1). We selected this medication because it is one of the two treatments that were compared against each other in SEAVUE, the target of our emulation and validation efforts.

We used the coefficients of the fitted placebo model to predict and remove the placebo-attributable component from the observed outcomes of these participants. The residuals from this process were interpreted as reflecting the adalimumab-attributable effect (Figure 1b). Across these patients the mean drug-attributable CDAI reduction was 68 points. We used these residuals to fit a second model for the adalimumab-attributable effect (Table 2).

As an exploratory analysis we reviewed the significant predictors of a response to adalimumab and compared these to the corresponding results from the placebo model. Although the sample size was relatively small, we noted a strong signal for age as a negative predictor: additional decades of life were associated with an 18-point reduction in the response to adalimumab. Interestingly, the direction of this effect was the opposite of that seen in the placebo-only model, suggesting that this coefficient might not have been identified as significant had it not been handled as an interaction term as we did.

### EXTERNAL VALIDATION

To validate our method, we designed an *in-silico* study to emulate SEAVUE, the only head-to-head study of FDA-approved biologics for Crohn’s disease to date^3^. In SEAVUE, biologic-naive patients with active Crohn’s disease were randomly to receive either adalimumab or ustekinumab as treatment. The primary endpoint was clinical remission at week 52, defined as a CDAI less than 150. Secondary endpoints included clinical remission at the time of all study visits, including week eight.

We identified all participants from the three ustekinumab-related trials who were biologic-naive. We identified 149 subjects who were assigned to ustekinumab and 135 participants assigned to placebo. We noted that the observed responses of the 135 placebo recipients reflected a combination of individual-specific variability and trial-specific variability (Figure 3). We therefore reasoned that to simulate the effect of treatment assignment, we needed to ‘add back’ the conditional mean effect associated with adalimumab to the outcomes of the placebo recipients (Figure 1c). Using the model coefficients identified in the adalimumab-attributable regression model (Table 2), we computed and added this extra reduction in the CDAI to the observed week eight outcomes of the placebo cohort.

Finally, we computed the proportion of patients who were in clinical remission at week eight, comparing the results of the observed ustekinumab recipients with that of the patients simulated to have received adalimumab and subject to the same background placebo effect (Figure 1d). We found that ustekinumab and adalimumab appeared to be equally efficacious, with 45% and 46% of the cohorts in remission. This result closely matched that of SEAVUE (p=0·9), which found 50% and 48% of these corresponding cohorts in remission (Table 3). Our simulated trial was similar in sample size to SEAVUE, with 149 and 135 patients receiving ustekinumab and adalimumab in our study, compared to 191 and 195 in SEAVUE.

**Table 3:** Clinical remission results at week 8.

|  | Ustekinumab (UST)<br>n/N (%) | Adalimumab (ADA)<br>n/N (%) | p-value |
| --- | --- | --- | --- |
| Published SEAVUE Results | 96/191 (50.3) | 93/195 (47.7) | - |
| Predicted SEAVUE Results, Primary Analysis | 67/149 (44.9) | 62/135 (45.9) | 0.905 |
| Sensitivity Analysis: Trials with High Capture of Outcomes | 67/149 (44.9) | 60/135 (44.4) | 1.000 |
| Sensitivity Analysis: Complete Cases | 66/148 (44.6) | 65/128 (50.3) | 0.398 |
| Sensitivity Analysis: Information Leakage | 67/149 (44.9) | 67/135 (49.6) | 0.475 |
| Negative Control, No Normalization | 67/149 (44.9) | 119/239 (49.8) | 0.403 |

We tested the robustness of this result using three sensitivity analyses. In the first we removed two trials (PRECISE1, ENACT) associated with the greatest degree of outcome missing data (Supplemental Figure 3). In the second, we performed a complete case analysis (deleted patient data associated with missing outcomes) as an alternative to last-observation-carried-forward imputation. In the third we removed all participant data emanating from an ustekinumab trial from the placebo training data, to address a possibility of information leakage. Our results remained unchanged over all sensitivity analyses (Table 3), supporting the robustness of our primary findings as well as the validity of our overall methodology.

Finally, we sought to evaluate the value of using our modeling approach compared to a simpler approach using published trial results. One barrier we noted to the latter was that the aggregated response of the TNFi-naive subcohorts at week eight was only published in one out of the six trials that we included for this comparison of ustekinumab and adalimumab, making it impossible to emulate SEAVUE using this approach. Separate from this, and to specifically evaluate the value of normalizing disparate cohorts using placebo models, we simulated the potential results of our head-to-head assessment without a normalization step. Under this scenario, the unnormalized adalimumab cohort response was 50% (Table 3). While this was not statistically significant compared to the observed ustekinumab response of 45% (p=0·4), it reflects a trend towards a difference. We interpreted this as reflecting a degree of bias that could plausibly result in false positives in other similar studies, but one that is analytically controllable using our method.

## DISCUSSION

We developed a new method for meta-analyzing individual participant data (IPD) from heterogenous randomized trials lacking a shared control group. We validated our methodology by successfully reproducing a major endpoint of SEAVUE, a recent head-to-head trial of biologic therapies in Crohn’s disease^3^. Our method involved several steps: identifying and isolating parallel arm cohorts from the available trials, harmonization and quality control, separately modeling the placebo effect from drug-attributable effects, and sequentially partitioning and assembling different sources of variation to accurately simulate the outcomes of a suitably normalized comparator group.

After decades of calls for greater data sharing^14–16^ we are now seeing many new platforms for accessing clinical trials data. The availability of these data has opened opportunities for researchers to verify published results as well as answer new questions using these data. This has never been more important, with the cost of new phase 3 clinical trials current at $20M and climbing^17^.

Although the growing availability of IPD portends well for the future of research, it has revealed new analytical challenges that require new methods. Existing methods for conducting IPD meta-analyses typically involve including trials with near-identical study designs, including fully parallel-design cohorts and shared placebo comparator arms. When these criteria are not met, problematic trials are often excluded from a given meta-analysis, sometimes in subtle ways. This substantially limits the numbers of questions that might already be answerable using existing clinical data. In some cases, this common practice might even introduce bias.

This work suggests that there may be better ways to handle this heterogeneity and discover new and trustworthy signals from these data. This method as well as extensions therein may substantially increase the numbers of studies that can be done, uncovering new evidence on comparative efficacy, safety, and ultimately precision medicine. Taking the example of Crohn’s disease, a major motivation for conducting the SEAVUE trial is the current level of uncertainty regarding the comparative efficacy of already approved treatments. Methods such as what we propose here can address these gaps, particularly as more therapies are approved and thus the number of potential head-to-head comparisons grows exponentially.

While we have illustrated this methodology in a comparative efficacy analysis, this approach may have significant value in other contexts. Models for the placebo effect, such as we demonstrate here, may help improve the design and statistical power of clinical trials across diseases. Moreover, the use of cohort normalization methods may be useful to improve the robustness of external control arm studies. These are studies that typically utilize real-world data to draw indirect inferences against controlled cohorts, typically single-arm intervention studies. However, our analysis suggests that a major driver of the large placebo effects in Crohn’s disease is the large unmodeled variation in the CDAI. Future work is needed to improve the measurement of Crohn’s disease activity.

We acknowledge several limitations. First, although we undertook extensive efforts to harmonize the data, we could not perfectly reproduce all covariate statistics as published. It is likely that we could have overcome these issues with access to the original analytical code. Nonetheless, the degree of deviations from published results was small, and our primary results remained robust to many sensitivity analyses. Future efforts involving pre-harmonization to a common data model may improve the reproducibility and feasibility of these IPD meta-analyses. Second, we were unable to include many important covariates like race and ethnicity. Most included studies did not capture ethnicity. Some studies did capture race but showed evidence of significant skew towards white participants. This likely reflects the historical underrecognition of the importance of these factors. Lastly, we note that our validation was somewhat underpowered and was performed in the context of just one disease. This is largely a function of the relative rarity of clinical trials (the source of our data and sample size), and especially head-to-head trials like SEAVUE. This underscores the importance of methods for learning more from these small but high-quality data. Future studies are needed to confirm the robustness and generalizability of our methodology to other diseases.

In conclusion, we developed a new method for meta-analyzing data from heterogeneous trials lacking a common control group. We validated this method by reproducing the results of a recent comparative efficacy trial using pre-existing data. We are sharing our code for others to replicate and build upon these methods, and ultimately uncover new insights using the data we already have.

## Data Availability

The raw data are owned by the aforementioned trial sponsors. The data may be accessed for reproduction and extension of this work following an application on the YODA and Vivli platforms and execution of a data use agreement. The analytical code is available as supplemental data files accompanying this manuscript.

## SUPPLEMENTAL METHODS

### QUALITY CONTROL, HARMONIZATION, MISSING DATA

We performed extensive quality control evaluations of the included trials and data (Figure 1a). This included confirming our ability to reproduce published statistics on the trial cohorts at baseline as well as the study primary endpoint (Supplemental Figure 3). We were able to exactly reproduce most of the study results. Where discrepancies occurred, they were generally minor and fell within a 10% error bound. We reported major discrepancies to the study sponsor as per agreement. We attempted to completely eliminate all discrepancies, but this was not possible due a variety of factors, including lack of access to the original analytic code or the complete analytic dataset, and inability to contact the original analysts.

We completed an assessment of data availability for all study variables (Supplemental Figure 4). Target variables included demographic features, CDAI at baseline and week eight, baseline inflammatory biomarkers, concomitant steroid and immunomodulator use, history of treatment with tumor necrosis factor-alpha inhibitors (TNFis), and other disease-related features. We identified nine variables that were universally available across all trials and thus could be used for downstream modeling: Age, Sex, BMI, baseline CDAI, c-reactive protein (CRP), history of TNFi use, oral steroid use, immunomodulator use, and ileal involvement.

Only 3% of the participants had at least one missing covariate at baseline. Continuous variables were addressed by median imputation, and participants with missing categorical variables were dropped from the dataset (N=86). 11% of the participants had a missing value for the outcome at week eight. To handle this, we used last-observation-carried-forward to impute these values, typically using measurements from week six and four. This is the typical practice for the analysis of these trials in regulatory submissions and was the prespecified approach in the protocols for all included trials. The variable corresponding to a history of TNFi use was available in all recent trials that occurred after the approval of the very first TNFi medication. Older trials of the first TNFis commonly excluded patients who had a history of exposure to other drugs from this class but did not include this feature as an actual variable in the data set. In these cases, we deterministically imputed this variable corresponding to no prior use.

Other variables of a priori importance could not be included in this study. Ethnicity was not collected in most trials. Race was missing in some trials, but when it was captured, it reflected significant imbalance (88% of participants were white). Other disease specific variables such as disease behavior and duration were also not uniformly captured across studies and thus could not be included in this meta-analysis.

### STATISTICAL COMPUTING

Programming was performed in the R language, using the packages *dplyr*^18^, *lme4*^19^, *lmerTest*^20^, *data*.*table*^21^, *ggplot2*^22^, *ggpubr*^23^, *sjstats*^24^, *patchwork*^25^, and *gridExtra*^26^. The analytical code was independently reviewed by a second member of the team.

### MODEL FOR ESTIMATING THE PLACEBO EFFECT

We fit a linear mixed effect model to predict the placebo effect on each patient’s CDAI reduction at week 8. The model was trained on the placebo arms of the six placebo-controlled trials. We denote them as trial 1 to trial 6 to simplify the notation. The CDAI reduction of patient *j* from trial *i* in the placebo arm at week 8 is denoted as 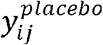 and assumed to be related to the nine predictors *D*_*ij*,1_, …, *D*_*ij*,9_,, the centered study year *T*_*i*_, and the trial-specific random effect *S*_*i*_ as in the following equation:

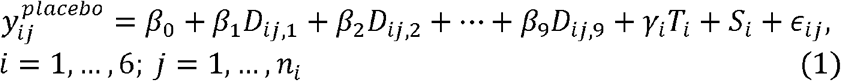

Where 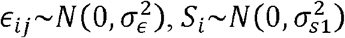, and *n*_*i*_ is the sample size of each trial, respectively.

### MODEL FOR ESTIMATING ADALIMUMAB DRUG-ATTRIBUTABLE EFFECT

After fitting the placebo-effect model, we used the coefficients of model (1) to predict the placebo-attributable component of the observed outcomes of the participants from three study cohorts assigned to receive adalimumab at the FDA-approved dose for treatment induction. We name them as trial 7 to trial 9 to simplify the notations. Denoting the observed CDAI reduction at week 8 of patient *j* from trial *i* as *y*_*ij*_ and the predicted placebo-attributable component as 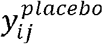, we assume the difference 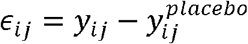 reflects the adalimumab drug-attributable effect and is related to the same nine predictors and trial-specific random effect of each adalimumab trial as in the equation below:

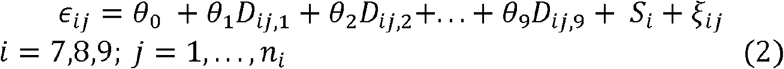

Where 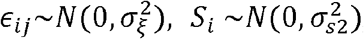, and *n*_*i*_ is the sample size of each trial, respectively.

### MODEL FOR EXTERNAL VALIDATION

To emulate SEAVUE, we identified all placebo-arm participants from the three ustekinumab-related trials who were biologic-naive as the simulated adalimumab cohort. The observed CDAI reduction of the participants at week 8 are denoted as 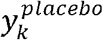, where *k* = 1, …, 135. We then use the coefficients of model (2) to predict the adalimumab drug-attributable effect of the simulated cohort and denote it as *ê*_*k*_. The CDAI reduction at week 8 of each simulated adalimumab participant is calculated by 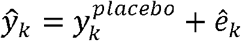. The number of remission *N*_*rem*_ is calculated by the count of *BaselineCDAI*_*k*_ − *ŷ*_*k*_ ≤ 150. The remission rate is calculated by *N*_*rem*_/135.

### CONTRIBUTORS

VAR conceived the study and obtained access to the data. SW, VGR, DVA, and VAR designed the study, analyzed the data, and drafted the manuscript. AM performed the risk of bias assessment. SW performed an independent review of the analytical code. All authors interpreted the data and critically edited the manuscript.

## ACKNOWLEDGEMENTS

The authors thank all the trial sponsors (Johnson & Johnson, AbbVie, UCB, Takeda, Biogen) for contributing the necessary participant-level data to carry out this study, and the participants of these studies consenting for their deidentified data to be shared in this way. They thank both the Yale Open Data Access Project and Vivli for enabling data access on a single platform. They thank Isabel Elaine Allen, Chiung-Yu Huang, and Mi-Ok Kim for biostatistical advice. They thank Cong Zhou for her contributions to earlier versions of this work.

This study, carried out under YODA Project #2018-3476, used data obtained from the Yale University Open Data Access Project, which has an agreement with JANSSEN RESEARCH & DEVELOPMENT, L.L.C.. The interpretation and reporting of research using this data are solely the responsibility of the authors and does not necessarily represent the official views of the Yale University Open Data Access Project or JANSSEN RESEARCH & DEVELOPMENT, L.L.C.

This publication is based on research using data from data contributors Johnson & Johnson, AbbVie, UCB, Takeda, Biogen that has been made available through Vivli, Inc. Vivli has not contributed to or approved, and is not in any way responsible for, the contents of this publication

